# Rapid establishment of public sector COVID-19 test and treatment programs across seven low- and middle-income countries: Implementation strategies and program monitoring results

**DOI:** 10.1101/2024.11.21.24317710

**Authors:** Caroline E. Boeke, Nervine Hamza, Chukwuemeka Agwuocha, Okechukwu Amako, Khamsay Detluexay, Michelle Gao, Bridget C. Griffith, Yamikani Gumulira, Elina Urli Hodges, Jessica Joseph, Norman Lufesi, Emily Macharia, Nyuma Mbewe, Elizabeth McCarthy, Faustina O. Mintah, Moses Mukiibi, Mwaba Mulenga, Alida Ngwije, Lawrence Ofori-Boadu, Ijeoma Uzondu Okoli, Sompasong Phongphila, Christian Ramers, Sean Regan, Evarist Twinomujuni, Edson Rwagasore, Jessica Tebor, Krishna Udayakumar, the COVID-19 Treatment QuickStart Consortium

## Abstract

The COVID-19 Treatment QuickStart Consortium[1] worked with governments in 7 low- and middle-income countries, Ghana, Laos, Malawi, Nigeria, Rwanda, Uganda, and Zambia, to implement COVID-19 test-and-treat programs at 776 health facilities, including training over 5,000 staff and facilitating a donation of 11,300 courses of the oral antiviral nirmatrelvir/ritonavir for treatment. This paper describes the process of implementing COVID-19 test-and-treat programs in each country, provides aggregate program monitoring data on numbers tested and treated from program initiation through June 2024, and analyzes program enablers and challenges. Between country-level program initiation (ranging from December 2022 for Zambia to September 2023 for Uganda) and June 2024, a total of 731,970 SARS-CoV-2 tests were conducted. Of 6,724 positive tests, a subset were documented to meet eligibility criteria for nirmatrelvir/ritonavir initiation, and 3,041 patients were prescribed nirmatrelvir/ritonavir. The largest number of prescriptions was in Zambia. Program enablers included decentralization of services, task-shifting from higher to lower health worker cadres, increased access to point of care antigen tests - including self-tests, and the integration of COVID-19 with other health services. Challenges included COVID-19 de-prioritization at the time of program rollout, test commodity stockouts and expiries, and dwindling national surveillance efforts. Learnings from rapid initiation and scale-up of COVID-19 test-and-treat programs in these 7 countries can be used to inform future pandemic preparedness strategies in LMICs.

## Introduction

The COVID-19 pandemic has led to over 9 million deaths globally as of June 2024.[2] Effective vaccines[3] and treatments[4] have helped prevent a higher death toll. However, over the course of the pandemic, low- and middle-income countries (LMIC) experienced delayed and more limited access to diagnostics, vaccines, and treatment.

In particular, nirmatrelvir/ritonavir, an oral antiviral demonstrated to reduce risk of hospitalization and death due to COVID-19,[4] was made available in high income countries after US FDA Emergency Use Authorization in December 2021, but the first reported delivery to LMIC by global buyers did not take place for another 15 months.[5, 6] Nirmatrelvir/ritonavir is intended for use through a test-and-treat model, in which individuals who test positive for COVID-19, have a mild/moderate infection, and are defined as high-risk (including older individuals or those with co-morbidities such as HIV, hypertension, cardiac disease, diabetes) are initiated on treatment within 5 days of symptom onset. Treatment with this 5-day regimen in this group of individuals reduces the risk of progression to severe/critical COVID-19, mitigating hospitalizations and death.[7–10] Limited data is available in LMIC contexts on real-world COVID-19 test-and-treat program implementation and utilization of nirmatrelvir/ritonavir.

The COVID-19 Treatment QuickStart Consortium worked with governments in 7 LMIC (Ghana, Laos, Malawi, Nigeria, Rwanda, Uganda, and Zambia) to develop and roll out COVID-19 test-and-treat programs from 2022-2024. Support included facilitating a nirmatrelvir/ritonavir donation as well as providing technical assistance on program implementation and monitoring and evaluation (M&E). An evaluation was conducted across the program countries; details on the evaluation methodology have been previously published.[11] This manuscript aims to describe program implementation approaches across the 7 countries, present aggregate M&E results across the 7 countries, highlight key program enablers and challenges, and facilitate sustainability planning for COVID-19 testing and treatment programs.

## Program Implementation Approach

### The QuickStart Consortium

The COVID-19 Treatment QuickStart Consortium, a partnership between Duke University, Americares, the Clinton Health Access Initiative, and the COVID Collaborative, received funding from Open Society Foundations, Pfizer, and the Conrad N. Hilton Foundation to introduce donated nirmatrelvir/ritonavir in 10 LMICs and provide technical assistance to governments in implementing COVID-19 test-and-treat programs. As of July 2024, 8 of the initial 10 countries had moved forward with approving the importation of the drug and implementing programs, and 7 had M&E data available.

### COVID-19 Test-and-Treat Program Approach

Broadly, implementation of test-and-treat programs for COVID-19 through the QuickStart program consisted of the following:

1. **Collaboration on guidelines development and implementation planning** including development of national policies, clinical guidelines, program rollout strategies, and national training programs, as well as generation of standard operating procedures (SOPs) for test operation and administration, results interpretation, quality assurance/quality control, treatment initiation, drug-drug interaction, and reporting.
2. **Supply and distribution planning for decentralized testing with point-of-care antigen (POC-Ag) tests** that have received emergency use authorization by stringent regulatory authorities (SRAs) or World Health Organization (WHO) Prequalification, and which are nationally approved for clinical use.
3. **Facilitating the donation of antiviral treatment** (nirmatrelvir/ritonavir) and in some countries, self-tests. Nirmatrelvir/ritonavir received emergency use authorization (and more recently, full US Food and Drug Administration approval) by SRAs, and which is nationally approved for clinical use. QuickStart provided support obtaining rapid regulatory waivers or approvals, and in-country importation.
4. **Training of healthcare workers on COVID-19 diagnostics and clinical care, and equipping health facilities to implement test-and-treat** including a training of trainers, support to cascaded training to health facilities, ongoing site mentorship, and support to facilities in mapping patient flow.
5. **Support to national monitoring and evaluation systems** including, 1) introduction of new facility registers to track patient-level information related to the individuals tested and treated through these programs, and 2) implementation of data management solutions to facilitate monitoring of test-and-treat for national program management and integration of data into existing country-level COVID-19 dashboards, as available. Paper-based or electronic registers were initiated at facilities which tracked data including patient characteristics, POC-Ag test and results, antiviral treatment eligibility, treatment regimen, treatment initiation dates and patient outcomes.
6. **Quality Control (QC) activities for post market surveillance,** supporting Ministries of Health (MOH) to ensure adequate pharmacovigilance systems in place to ensure reporting of serious adverse events (SAEs) associated with nirmatrelvir/ritonavir.
7. **Program advocacy at the community level** including development of information, education, and communication (IEC) materials aimed to educate community members on availability of test-and-treat at local facilities and eligibility criteria for treatment.

QuickStart was the first to establish COVID-19 test-and-treat programs in four countries (Laos, Nigeria, Uganda, Zambia); three countries (Ghana, Malawi, Rwanda) were conducting test-and-treat activities outside of QuickStart. Of note, some countries were also supporting COVID-19 test-and-treat work outside of the QuickStart program.

### Ethics Statement

The data presented in this manuscript came from aggregate sources rather than patient-level data; authors could not identify individual participants. However, this study was approved by the Duke University Institutional Review Board (Pro00111388). Consent was not obtained due to participant anonymity.

## Implementation Highlights by Country

Key characteristics of each country’s COVID-19 test-and-treat program are provided in Table 1. All facilities provided COVID-19 treatment free of charge. Most facilities were public sector sites; however, some private facilities were included in Zambia and Nigeria’s programs which agreed to provide services free of charge. Specific country program details are provided below.

**Table 1.** Characteristics of National Public Sector COVID-19 Test and Treat Programs by Country, as of June 2024.

|  | Ghana | Laos | Malawi | Nigeria | Rwanda | Uganda | Zambia |
| --- | --- | --- | --- | --- | --- | --- | --- |
| **Program leadership and policies** |  |  |  |  |  |  |  |
| National program leadership | Institutional Care Division, Ghana Health Service | Department of Healthcare and Rehabilitation, Ministry of Health | Clinical Services Team, Curative & Medical Rehabilitation Services Directorate, Ministry of Health | Department of Hospital Services, Federal Ministry of Health and the Nigeria Centre for Disease Control and Prevention | Pandemic Division, Rwanda Biomedical Centre | Department of Clinical Services, Ministry of Health | Infectious Disease and Clinical Care Department, Ministry of Health |
| National clinical and testing guidelines available | Yes[12] | Yes[13] | Yes[14] | Yes[15, 16] | Yes[17] | Yes[18] | Yes[19] |
| **Procurement** |  |  |  |  |  |  |  |
| Originator nirmatrelvir/ritonavir registered in country | Yes | No | No | No | No | No | No |
| Generic nirmatrelvir/ritonavir registered in country (WHO prequalified) | No | Yes | Yes | No | No | No | No |
| Procurement of diagnostics | Both domestically procured (Global Fund) and donations from development partners | Both domestically procured and donations | Both domestically procured (Global Fund) and donations from partners | Both domestically procured (Global Fund) and donations from partners | Donated | Both domestically procured (Global Fund) and donations from partners | Both domestically procured (Global Fund) and donated by partners |
| Date first drugs arrived in country | March 2023 | February 2023 | March 2023 | April 2023 | February 2023 | June 2023 | December 2022 |
| Donation quantity through QuickStart program | 1000 | 1500 | 700 | 2,600 | 1,000 | 500 | 4,000 |
| **Testing program** |  |  |  |  |  |  |  |
| Self-testing available | No | Yes | No | No | Previously available | No (But provided for in the revised testing guidelines) | No |
| Testing available outside of health facilities | No | Yes | Yes, but only for healthcare workers | Yes | Yes | Yes | Yes, by trained community-based volunteers |
| Mass testing campaigns conducted by government | No | Yes | Yes, as a study in 2022 | Yes | Yes | No | No |
| Bi-directional testing | Yes in some facilities. ILI/C-19 and TB/C-19 | Yes (with ILI/SARI and TB) | No | Yes, Malaria/C-19, HIV/C-19, TB/C-19 | Yes | No | Yes, TB/C-19 |
| Test cost to users | Free | Free | Free | Free | Free | Free | Free |
| **National program management** |  |  |  |  |  |  |  |
| Stock management system / process | Push distribution mechanism using the last mile distribution cycle from the Supply, Stores and Drug Management Division of Ghana Health Service to regional medical stores | National eLMIS (mSupply system) used at all warehouse levels to manage stock. Department of Healthcare and Rehabilitation creates distribution plan for treatment; National Center of Laboratory and Epidemiology manages test distribution. The Medical Product and Supply Center oversees stock management and distribution | Test kits: pull system used and the diagnostic department are responsible for distribution<br>Treatment: push system used based on consumption data. The national PSM manages the distribution | Push distribution strategy from national Central Medical Store to states; consumption reports used to monitor supply | Distributed to Rwanda Biomedical Supply (RMS) branches based at each district to serve health facilities in catchment area. Hospitals hold treatment and deliver to health centers when there are positive patients. | Quantification and Procurement Unit (QPPU) of the Department of Pharmaceuticals and Natural Medicines (DPNM) Ministry of health and partners, and distributed to facilities based on regional disease burden by Central Public Health Laboratory (CPHL) using allocation lists | Managed by Zambia Medicines and Medical Supplies Agency. Facilities determine needs and order test kits using electronic logistics management information system (eLMIS). Drugs distributed using a push system |
| Data system | Paper-based; hospital-based electronic medical record in some facilities | Electronic - Google form | Paper-based; currently working on electronic reporting | Diagnostic data: electronic – Surveillance Outbreak Response Management and Analysis System (SORMAS). Treatment data: paper-based; electronic medical records in some health facilities | Electronic – DHIS2 patient tracker; paper-based tracker at hub facilities. Improved data reporting coverage from January 2024 onwards. | Diagnostic data: Electronic – proprietary system (Laboratory Results Delivery System). Treatment data: Paper based | Electronic Integrated Disease Surveillance and Response (eIDSR) system, REDCap, SmartCare and paper-based registers and clinical forms |
| National HMIS now includes C-19 antivirals | No | No | No | Yes | Yes | No | No |

### Ghana

Ghana has documented 172,075 COVID-19 cases and 1,462 deaths from 2020 through April 2024, with three noticeable waves from May-August 2020, January-March 2021 and July-September 2021.[20] Of note, data from POC-Ag tests, which have been offered in all facilities since 2022, are not yet captured in national surveillance data. SARS-CoV-2 vaccines were introduced in March 2021. The COVID-19 Treatment QuickStart program is managed through the Ghana Health Service-Institutional Care Division. Leveraging available POC-Ag tests and a donation of 1,000 treatment courses through QuickStart, the first test and treat facility was activated – including staff members trained and test-and-treat commodities available for patients - in May 2023. Since program initiation, through QuickStart, 154 healthcare workers (doctors/physician assistants, nurses, pharmacists, medical laboratory scientists, disease control officers, health promotion officers) were trained across 16 health facilities (primary, secondary and tertiary level hospitals) across 5 regions using a Training of Trainers approach. With support from USAID/RISE and The Aurum Institute, Ghana Health Service activated additional facilities, increasing total coverage to approximately 54 facilities across 11 of the 16 regions in Ghana. Entry points for screening are inpatient wards, outpatient clinics, and emergency rooms.

### Laos

Laos has documented approximately 207,300 cases of COVID-19 and 750 deaths[21] as of July 2024, with notable peaks in August 2021 and January 2022. COVID-19 vaccines first arrived in Laos in November 2020, becoming available to the public in the first quarter of 2021. The national COVID-19 test and treat program is managed by the Department of Healthcare and Rehabilitation within the Ministry of Health. Unitaid also provided complementary program support in Laos. Laos received its first donation of nirmatrelvir/ritonavir courses in February 2023; 1,500 courses were donated through QuickStart and the first facility was activated in July 2023. Since program initiation, 238 providers have been trained, and 37 facilities, including central, provincial, and district hospitals, offer treatment across all 18 provinces. The entry points for screening, testing, and linkage to treatment typically occur in emergency rooms and outpatient departments. Key partners supporting the program include the WHO and UNICEF.

### Malawi

Malawi has documented 89,564 cases of COVID-19 and 2,687 deaths as of June 2024,[22] with peaks in January 2021, July 2021, January 2022 and January 2024. COVID-19 vaccination was officially launched in March 2021. The COVID-19 test and treat program is led by the Curative and Medical Rehabilitation Directorate of the Ministry of Health. 700 treatment courses were donated through QuickStart in March 2023 and the first facility was activated in July 2023. 12 health facilities across all 3 regions were activated to provide test and treat services. 18 additional facilities were activated through the EPIC program funded by USAID and supported by FHI360. Since the program’s inception, 1554 providers from the 12 QuickStart facilities received training. The entry points for screening, testing and treatment include HIV antiretroviral therapy clinic, non- communicable disease, and outpatient clinics/departments. FHI360 is the only additional partner actively involved in the implementation of COVID-19 test and treat programs in Malawi.

### Nigeria

Nigeria documented 266,313 cases of COVID-19 and 3,155 deaths as of February 2023,[23] with peaks from December 2020-February 2021, December 2021-February 2022, and December 2022- February 2023. The COVID-19 pandemic response in Nigeria has been driven by the Department of Hospital Services of the Federal Ministry of Health and the Nigeria Centre for Disease Control. Case Management is domiciled in the Department of Hospital Services of the Federal Ministry of Health while the Nigeria Centre for Disease Control which manages diagnostics and other surveillance strategies. Nigeria received the first shipment of the COVID-19 vaccines through the COVAX Facility in March 2021. 2,600 treatment courses were donated through QuickStart, with the first treatment facility activated in June 2023. 1,775 healthcare workers were trained across 69 primary and secondary health facilities in 3 high-burden states, Rivers, Nasarawa, and Ogun, as well as the Federal Capital Territory. Five private facilities were selected to demonstrate the feasibility of the test and treat approach in private facilities. Entry points included outpatient, HIV treatment, and inpatient clinics, and community facilities. Bidirectional testing was also employed, focused on the integration of COVID-19 testing with testing for other diseases such as those seen for fever, suspected of malaria, or presenting for routine immunization services, and in febrile patients. Other key partners include Institute of Human Virology of Nigeria and the AIDS Prevention Initiative in Nigeria. Partners improved case detection through testing across different populations such as people living with HIV and tuberculosis.

### Rwanda

Rwanda has documented 132,489 cases of COVID-19 and 1,466 deaths as of May 2024[24], with peaks in COVID-19 cases from December 2020 to February 2021. The country introduced and scaled antigen testing for SARS-CoV-2 in January 2021 and COVID-19 vaccines in March 2021.

The COVID-19 test and treat program is managed through Public Health Surveillance and Emergency Preparedness and Response at the Rwanda Biomedical Center, an implementation agency of the Ministry of Health. The first oral antivirals arrived in the country in June 2022 (procured bilaterally between the government and Pfizer), followed by additional treatment courses received through QuickStart in February 2023; a total of 1,000 treatment courses were donated through QuickStart and the first facility was activated in March 2023. Since program initiation, an estimated 895 providers (medical doctors and nurses) have been trained and 589 hospitals and health centers offered COVID-19 test and treat services across the country. The primary criteria to treat patients per Rwanda’s national guidelines is based on presenting within 5 days of symptom onset and having mild/moderate symptoms. Entry points at facilities include outpatient departments and the laboratory for inpatients. Rwanda also has conducted several mass screening campaigns in the community. Other key partners supporting COVID-19 test and treat include USAID/RISE through Jhpiego which activated 8 of the 50 hospitals in the country.

### Uganda

Uganda has documented approximately 170,800 cases of COVID-19 and 3,650 deaths as of May 2024,[25, 26] with peaks in December 2020, June 2021, July 2022, and December 2022. The first batch of COVID-19 vaccines was received in March 2021 via the COVAX Facility. The national COVID-19 test and treat program is managed through the Clinical Services Department within the Ministry of Health. 500 treatment courses were donated through QuickStart and the first QuickStart supported facility was activated in September 2023. Since the program’s initiation, 284 providers received training and 17 hospitals have offered test and treat programs across 14 of the 15 regions in the country. Entry points at facilities include screening in outpatient wards, special clinics and medical in-patient wards; sample collection often took place in designated locations next to the laboratory. Other key partners supporting COVID-19 test and treat include the Infectious Diseases Institute that supported last mile delivery and pharmacovigilance of the donated nirmatrelvir/ritonavir.

### Zambia

Zambia has recorded approximately 354,126 cases of COVID-19 and 4,079 deaths as of June 2024[27]. The highest peaks in cases were observed during January 2021, June 2021 and January 2022. The first batch of vaccines was received in April 2021, via the COVAX Facility. The national COVID-19 test and treat program is managed through the Directorate of Infectious Diseases as well as the Department of Clinical Care within the Ministry of Health with support from the Zambia National Public Health Institute (ZNPHI). 4,000 treatment courses were donated through QuickStart; the first QuickStart-supported test and treat facility was activated in December 2022 (the first of the entire program). Since program initiation, 27 national level trainers, over 133 facility healthcare providers, and 103 community-based volunteers have been trained, and 36 health facilities activated to offer test and treat services across 8 provinces in the country. Two private sector facilities were included; the remainder were public sector facilities. COVID-19 test and treat entry points at facilities included inpatient departments as well as out-patient departments such as emergency, HIV clinics, and TB clinics. Other key partners supporting COVID-19 test and treat include ICAP at Columbia and Centre for Infectious Disease and Research in Zambia.

## COVID-19 QuickStart Test and Treat Program Facility Characteristics

Table 2 provides an overview of the facilities implementing test and treat programs through the QuickStart program. Data were gathered through facility information forms administered at facilities or through an online portal. In summary, test and treatment programs were implemented at 776 facilities; 238 facilities with QuickStart COVID-19 test-and-treat programs had facility information data available (in Rwanda, most health centers did not submit facility data). 66.1% of these facilities were hospitals; the remainder were health centers or clinics. 72.0% were located in urban or semi-urban areas. 33.5% were supported by partner organizations in addition to the government. 72.4% were open for COVID-19 testing and treatment 7 days per week. At the time of data collection, 35.6% offered PCR testing, 65.7% offered antigen testing, and 8.9% offered self- testing kits for personal use. COVID-19 screening was most commonly offered in outpatient departments (70.3% of facilities), inpatients (61.4%), the emergency department (60.2%), TB clinics (53.9%), HIV clinics (51.3%), and non-communicable disease clinics (51.3%). COVID-19 oral antiviral treatment was most commonly offered in outpatient departments (76.3%) followed by inpatient departments (55.9%).

**Table 2.** Characteristics of QuickStart-supported Facilities offering COVID-19 Test and Treat Programs.

|  | Ghana | Laos | Malawi | Nigeria | Rwanda | Uganda | Zambia |
| --- | --- | --- | --- | --- | --- | --- | --- |
| Number of QuickStart facilities activated and implementing test and treat as of June 2024 | 16 | 37 | 12 | 69 | 589 | 17 | 36 |
| Facilities included in analysis below | 16 | 37 | 12 | 42<sup>1</sup> | 93<sup>1</sup> | 17 | 21<sup>1</sup> |
| Date of first facility activation | May 2023 | July 2023 | July 2023 | June 2023 | March 2023 | September 2023 | December 2022 |
| Geographic coverage of facilities (e.g., # of regions) | 5 of 16 regions | All 17 provinces and capital | All 3 regions | 3 of 36 states and capital | All public health facilities in country | 14 of 15 regions | 8 out of 10 provinces |
| Total number of staff trained on test and treat | 154 | 238 | 1554 | 1,775 | 895 | 284 | 133 |
| Facility type |  |  |  |  |  |  |  |
| Hospital | 100.0% | 89.2% | 50.0% | 64.3% | 44.1% | 100.0% | 85.7% |
| Health Center | 0.0% | 10.8% | 50.0% | 31.0% | 55.9% | 0.0% | 14.3% |
| Clinic | 0.0% | 0.0% | 0.0% | 4.8% | 0.0% | 0.0% | 0.0% |
| Facility location |  |  |  |  |  |  |  |
| Urban | 56.3% | 89.2% | 75.0% | 52.4% | 20.4% | 82.4% | 85.7% |
| Semi-urban | 43.8% | 10.8% | 8.3% | 31.0% | 18.3% | 17.6% | 14.3% |
| Rural | 0.0% | 0.0% | 16.7% | 16.7% | 57.0% | 0.0% | 0.0% |
| Very remote | 0.0% | 0.0% | 0.0% | 0.0% | 4.3% | 0.0% | 0.0% |
| Facility was supported by groups in addition to MOH | 18.8% | 18.9% | 25.0% | 38.1% | 26.9% | 41.2% | 28.6% |
| Facility offered COVID-19 testing and treatment 7 days per week | 81.3% | 91.9% | 16.7% | 23.8% | 86.0% | 82.4% | 95.2% |
| Types of testing available at facilities (select all that apply) |  |  |  |  |  |  |  |
| PCR | 81.3% | 27.0% | 50.0% | 21.4% | 5.4% | 88.2% | 57.1% |
| Antigen | 100.0% | 94.6% | 100.0% | 0.0% | 100.0% | 82.4% | 100.0% |
| Self-testing | 0.0% | 0.0% | 25.0% | 0.0% | 19.4% | 0.0% | 0.0% |
| Where patients are tested for C-19 at facilities (select all that apply) |  |  |  |  |  |  |  |
| Inpatient | 56.3% | 37.8% | 16.7% | 31.0% | 63.4% | 58.8% | 71.4% |
| Outpatient | 43.8% | 56.8% | 58.3% | 42.9% | 87.1% | 76.5% | 95.2% |
| Emergency | 50.0% | 45.9% | 16.7% | 23.8% | 48.4% | 41.2% | 76.2% |
| HIV clinic | 18.8% | 0.0% | 33.3% | 35.7% | 32.3% | 11.8% | 38.1% |
| TB clinic | 25.0% | 8.1% | 16.7% | 31.0% | 32.3% | 23.5% | 52.4% |
| Non-communicable disease clinic | 93.8% | 2.7% | 16.7% | 0.0% | 38.7% | 17.6% | 14.3% |
| Other | 25.0% | 21.6% | 66.7% | 54.8% | 7.5% | 35.3% | 23.8% |
| Where patients are initiated on C-19 treatment at facilities (select all that apply) |  |  |  |  |  |  |  |
| Inpatient | 81.3% | 51.4% | 0.0% | 45.2% | 58.0% | 76.5% | 76.2% |
| Outpatient | 81.3% | 27.0% | 41.7% | 100.0% | 87.1% | 76.5% | 85.7% |
| Emergency | 68.8% | 5.4% | 0.0% | 38.1% | 40.9% | 35.3% | 47.6% |
| HIV clinic | 25.0% | 2.7% | 0.0% | 69.0% | 30.1% | 23.5% | 33.3% |
| TB clinic | 25.0% | 5.4% | 0.0% | 33.3% | 30.1% | 29.4% | 28.6% |
| Non-communicable disease clinic | 0.0% | 0.0% | 0.0% | 0.0% | 33.3% | 23.5% | 9.5% |
| Other | 31.3% | 48.6% | 58.3% | 11.9% | 0.0% | 41.2% | 9.5% |

<sup>1</sup>Data from some facilities in Zambia not available due to them being self-activated / not directly supported by the Clinton Health Access Initiative. Data not available from 27 facilities in Nigeria with lighter touch program support. Data from Rwanda was collected via a voluntary online survey distributed to all facilities, and therefore should be considered a convenience sample. 13 facilities nationwide offer PCR testing in Rwanda.

## Testing and Treatment Volumes

Table 3 provides aggregate monitoring and evaluation data from each country from program initiation through June 2024. Aggregate data were gathered through national data reporting systems (as described in Table 1) between July and November, 2024. Across the 7 countries, 731,970 people were tested for SARS-CoV-2 at activated test and treat facilities, with the highest volumes of testing occurring in Rwanda (442,228), Nigeria (193,486), and Zambia (38,426). There were 6,724 positive tests across countries, with the highest numbers of positives in Zambia (3,314), Rwanda (1,516), and Malawi (691). The percentage of people who tested positive varied dramatically across countries depending on testing strategy (targeted vs. mass screening campaigns) and volume. 1,965 people were recorded by test and treat program staff as eligible for treatment.

**Table 3.** COVID-19 Treatment Volumes Achieved in Public Sector Programs from QuickStart Program Initiation through June 2024.

|  | **Ghana** | **Laos** | **Malawi** | **Nigeria<sup>1</sup>** | **Rwanda** | **Uganda** | **Zambia** | **Total** |
| --- | --- | --- | --- | --- | --- | --- | --- | --- |
| Date first test-and-treat facility activated | May 2023 | July 2023 | June 2023 | June 2023 | March 2023 | September 2023 | December 2022 | - |
| Number of facilities offering test-and-treat | 16 | 37 | 12 | 69 | 589 | 17 | 36 | 776 |
| Number of tests conducted | 5,750 | 26,236 | 14,697 | 193,486 | 444,228 | 9,147 | 38,426 | 731,970 |
| Number of positive patients | 289 | 297 | 691 | 362 | 1,515 | 256 | 3,314 | 6,724 |
| Number of patients recorded as eligible<sup>2</sup> | 209 | 56 | 295 | 165 | 401 | 130 | 709 | 1,965 |
| Number of patients prescribed nirmatrelvir/ritonavir | 194 | 52 | 282 | 161 | 790 | 106 | 1,465 | 3,041 |

<sup>1</sup>Data in Nigeria is through April 2024 due to testing stockouts from May-June 2024.
<sup>2</sup>This included presenting within 5 days of symptom onset, mild or moderate disease, and in a high-risk population. Some facilities had limited documentation of eligibility criteria, particularly in Rwanda where per national guidelines, being in a high-risk population was not always required for prescription. Zambia did not collect this information for the first 3 months of the program.

Eligibility criteria were not consistently documented in the data across all countries, and in some circumstances, providers used clinical judgment to make prescription decisions, or eligibility was not initially documented; this explains why in some countries, such as Zambia, the number of prescriptions is greater than the number recorded in the registers as eligible for nirmatrelvir/ritonavir. Ultimately, 3,041 people were prescribed nirmatrelvir/ritonavir, with the largest numbers of prescriptions in Zambia (1,461), Rwanda (786), and Malawi (282).

Figure 1 shows the number of patients testing positive over time by country as well as the number treated over time by country. Treatment numbers were highly variable over time, depending on the number of people who tested positive.

**Figure 1.**
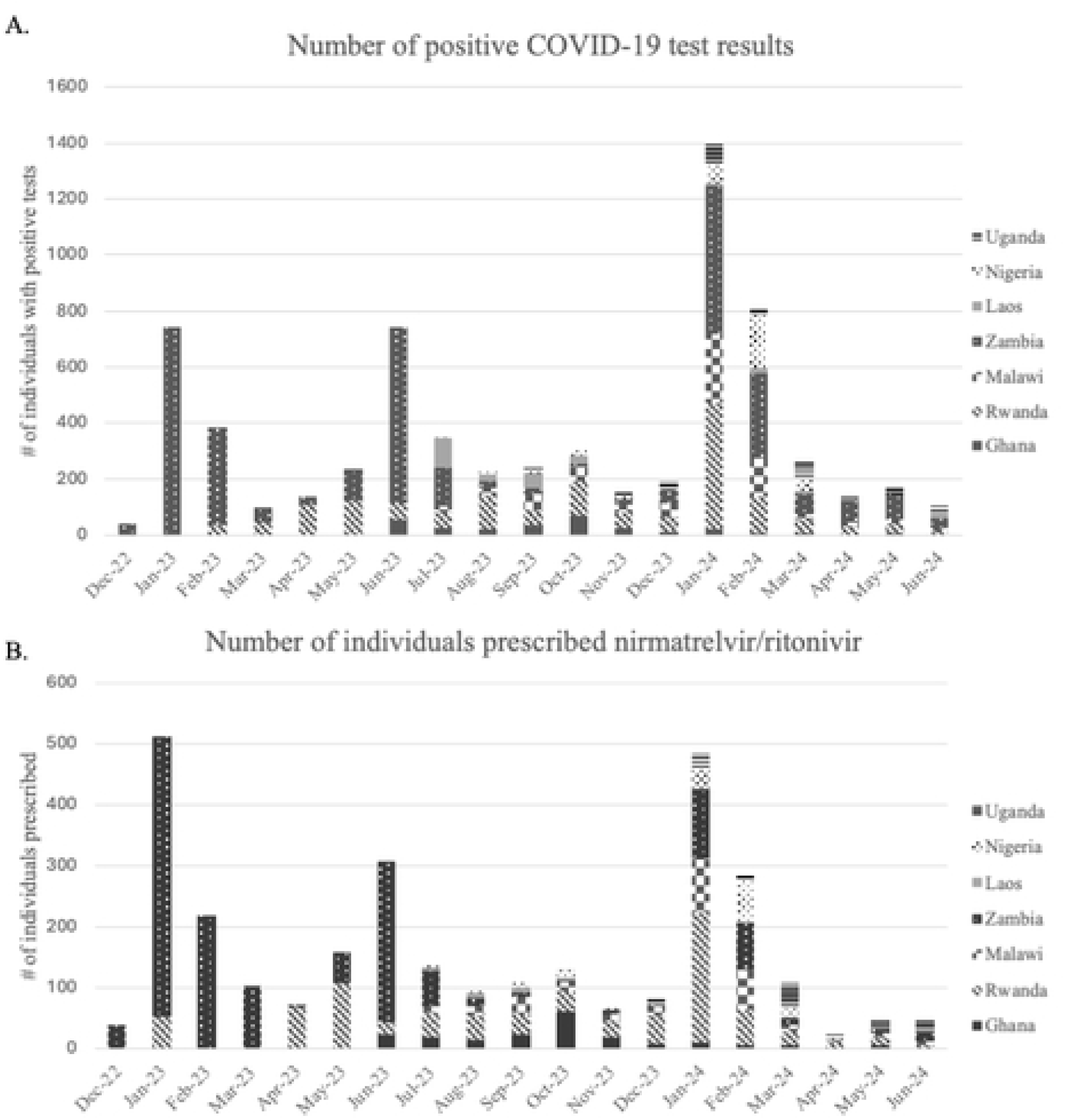
Number of A) positive SARS-CoV-2 tests per country over time and B) number of people treated with nirmatrelvir/ritonavir over time at activated QuickStart facilities

## Program Enablers

Key enablers of smooth program implementation across countries were identified based on communications with key stakeholders, and analysis of program data. First, decentralization of services to lower-level facilities and rapid task shifting to lower health worker cadres allowed for expanded access to services. This approach was more effective to facilitate rapid scale-up in comparison with limiting the provision of services to a small number of specialists. Secondly, access to POC-Ag tests, including self-testing in some country contexts, was another critical factor to facilitate rapid program scale-up. The investments made into scaling rapid diagnostic tests (RDT) before the QuickStart program launch were significant to facilitate a smooth rollout of treatment.

For example, the Ghana program leveraged previous experience activating COVID-19 antigen testing facilities across all regions of the country – prior to QuickStart and led by Ghana Health Service with support from the Clinton Health Access Initiative - to expedite test-and-treat program roll-out. Facilities were equipped with human resources and commodities to conduct testing, allowing training to extensively focus on clinical management. Third, integrated services delivery was critical to rapid implementation. Implementation worked best in facilities where there was a high degree of coordination between the clinical services, diagnostics, and supply chain departments to ensure smooth implementation. Integrated service delivery was less resource intensive and leveraged existing human resources and program structures, ultimately strengthening these structures. For example, Zambia leveraged TB and HIV infrastructure to set up the program leading to establishment of bi-directional screening of COVID-19 and tuberculosis, which ultimately led to better patient care for both disease areas.

In addition to these core factors, facility selection played a role in the ability to quickly launch the program in some contexts. For example, in Malawi, all health facility levels were considered; selection was based on regional disease burden, each facility’s ability to conduct testing, presence of doctors on site, and capacity to manage and store stock of drugs and tests, which ultimately led to the initial program rolling out smoothly at facilities best equipped to handle it. However, as noted above, decentralization of services to lower-level facilities is a critical longer-term goal.

Identification of champions to support with introduction and implementation of test and treat was another enabler. For example, in Zambia, there was strong Ministry of Health leadership through the case management specialist who championed the program and continued engagement on the part of the government, which sustained momentum. Finally, in some contexts such as Zambia, community engagement was viewed as an important enabler for program success.

## Challenges to Program Rollout

The greatest challenge highlighted by countries in implementing programs was COVID-19 fatigue by both communities and health care workers, as well as the WHO declaration ending the Public Health Emergency of International Concern designation in May 2023. With the reduced number of COVID-19 cases at the time of treatment scale-up, many did not see the importance of test and treat given the timing and other competing priorities. In particular, competing public health emergencies, including cholera in Malawi and Zambia, redirected attention from COVID-19. Relatedly, test commodity availability was a challenge across all countries. Once facilities stocked out of tests or the stock expired, it was challenging to revive testing efforts. For example, in 2022, with Global Fund funding, Ghana procured and supplied COVID-19 POC-Ag testing to all testing facilities across the country. The tests’ expiration dates occurred during implementation of the Quick Start program, causing significant stockouts across Quick Start supported facilities for a period of over two months (February to April 2024), which led to gaps in testing and treatment initiation. In Nigeria, there were also test stockouts from May to June 2024. Finally, national surveillance efforts dwindled during the period of program implementation, limiting availability to assess program progress. Without surveillance and testing, this reinforced the sentiment that COVID-19 was no longer a significant problem. Despite these challenges, these 7 countries did successfully roll out COVID-19 test and treat programs within a period of 12 months of donor support.

Two additional countries supported by QuickStart did not roll out the program within this timeframe, due to the impossibility to bring donated nirmatrelvir/ritonavir in country. In one country, South Africa, the drug was deemed not to be a cost-effective treatment for the public sector. In the other, Zimbabwe, delays in obtaining clearing documents in time to bring in drugs with adequate shelf-life led to an inability to advance the program. Finally, another country, Kenya experienced a delayed rollout of the program, meaning that treatment data were not available as of June 2024. Learnings from these challenges will be important to incorporate into planning for future pandemic preparedness response.

## Discussion

In summary, over a period of 19 months, 7 low- and middle-income countries (Ghana, Laos, Malawi, Nigeria, Rwanda, Uganda, and Zambia) were able to implement COVID-19 test-and-treat programs at 776 health facilities. This included training over 5,000 staff, facilitating a donation of 11,300 courses of nirmatrelvir/ritonavir for treatment, conducting 731,970 SARS-CoV-2 tests, and prescribing nirmatrelvir/ritonavir to 3,041 patients. Despite some challenges around de- prioritization of COVID-19 as a public health emergency, programs were able to leverage existing POC-Ag testing programs and approaches including decentralization, task shifting, and integration with existing services to rapidly roll out and scale up a test and treat program.

Limitations of this analysis include that testing and treatment data were gathered through country-level data systems and were in aggregate; therefore, there may be inaccuracies and incomplete data in some contexts, and data cannot be linked to track individual patients across the cascade of care. In addition, this analysis does not include patient outcome data. However, the study team plans to conduct additional analysis on individual-level patient registers in a subset of facilities/countries.

As the COVID-19 landscape evolves and the funding landscape becomes more constrained, many governments are updating their COVID-19 plans to determine response protocols for low-incidence periods and preparedness plans for future surges. This work includes strengthening surveillance systems, developing surge preparedness plans, putting mechanisms in place for increased medical countermeasures as needed, and identifying updated and integrated governance structures for continued program oversight. Learning from experiences rapidly rolling out COVID-19 test and treat programs will be critical to prepare for the next pandemic.

## Data Availability

Data available on reasonable request and approval of all relevant parties (e.g., country Ministry of Health).

## Acknowledgments

We acknowledge government partners and healthcare workers in QuickStart program countries for their dedication in implementing this program. We recognize Americares for their collaboration in this consortium as partners involved in the logistics and delivery of donated products. This work was funded by Pfizer, the Open Society Foundations, and the Conrad N. Hilton Foundation. We acknowledge all members of the COVID Treatment QuickStart Consortium: Krishna Udayakumar, Duke Global Health Innovation Center, Elina Urli Hodges, Duke Global Health Innovation Center, Ellen (Nellie) Bristol, Duke Global Health Innovation Center, Emily Macharia, Duke Global Health Innovation Center, Katharine Olson, Duke Global Health Innovation Center, Wenhui Mao, Duke Global Health Innovation Center, Cameron Wolfe, Duke University School of Medicine, Megan Oakes, Duke University School of Medicine, Sarah Gonzales, Duke University School of Medicine, Julie Miller, Duke University School of Medicine, Hayden Bosworth, Duke University School of Medicine, Joseph (Leo) Brothers, Duke Clinical Research Institute, Laine Thomas, Duke Clinical Research Institute, Monica Leyva, Duke Clinical Research Institute, Maria Grau-Sepulveda, Duke Clinical Research Institute, Sharon Stroud, Duke Clinical Research Institute, Ti Fadika, Duke Clinical Research Institute, Klay Shannon, Duke Clinical Research Institute, Suzanne Zeid, Duke Clinical Research Institute, Michael Merson, Duke Global Health Institute, Shanti Narayanasamy, Duke Global Health Institute Gary Edson, COVID Collaborative, Elizabeth McCarthy, Clinton Health Access Initiative, Sean Regan, Clinton Health Access Initiative, Jessica Tebor, Clinton Health Access Initiative Jessica Joseph, Clinton Health Access Initiative, Caroline Boeke, Clinton Health Access Initiative Nervine Hamza, Clinton Health Access Initiative, Bridget Griffith, Clinton Health Access Initiative, Alan Staple, Clinton Health Access Initiative, Fiona Gambanga, Clinton Health Access Initiative Hilda Shakwelele, Clinton Health Access Initiative, Alexander Martin-Odoom, Clinton Health Access Initiative, Alida Ngwije, Clinton Health Access Initiative, Chukwuemeka Agwuocha, Clinton Health Access Initiative, Ciru Wanjiru Ndichu, Clinton Health Access Initiative, Lorraine Kabunga, Clinton Health Access Initiative, Maame Nkansaa Asamoa-Amoakohene, Clinton Health Access Initiative, Mwaba Mulenga, Clinton Health Access Initiative, Philip Kimani, Clinton Health Access Initiative, Phyllis Chituku, Clinton Health Access Initiative, Prudence Haimbe, Clinton Health Access Initiative, Sompasong Phongphila, Clinton Health Access Initiative, Garrett Young, Clinton Health Access Initiative, Davis Karambi, Clinton Health Access Initiative, Tamara Mwenifumbo, Clinton Health Access Initiative, Vennie Nabitaka, Clinton Health Access Initiative, Vishal Brijlal, Clinton Health Access Initiative, Yamikani Gumulira, Clinton Health Access Initiative, Folu, Lufadeju, Clinton Health Access Initiative, Brenda Kateera, Clinton Health Access Initiative, Alexio Mangwiro, Clinton Health Access Initiative, Leslie Emegbuonye, Clinton Health Access Initiative, Christian Ramers, Clinton Health Access Initiative, Michelle Gao, Clinton Health Access Initiative, Okechukwu Amako, Clinton Health Access Initiative, Maria Corcorran, Clinton Health Access Initiative, Anthony Bless Dogbedo, Clinton Health Access Initiative, Faustina Ofosua Mintah, Clinton Health Access Initiative, Somsy Dongpadith, Clinton Health Access Initiative, Sabine Umuraza, Clinton Health Access Initiative, Rodrigue Ndayishimiye, Clinton Health Access Initiative Joseph Kalua, Clinton Health Access Initiative, Nere Otubu, Clinton Health Access Initiative, Simene Sangha, Clinton Health Access Initiative, Oluwatosin Akinwande, Clinton Health Access Initiative, Opeyemi Abudiore, Clinton Health Access Initiative, Evarist Twinomujuni, Clinton Health Access Initiative, Robert Kirungi, Clinton Health Access Initiative, Milimo Mweemba, Clinton Health Access Initiative, Megan Knox, Clinton Health Access Initiative, Khamsay Detleuxay, Department of Healthcare and Rehabilitation, Ministry of Health, Lao PDR, Bounxou Keohavong, Food and Drug Department, Ministry of Health, Lao PDR, Viengsakhone Louangpradith, Department of Healthcare and Rehabilitation, Ministry of Health, Lao PDR, Edson Rwagasore, Rwanda Biomedical Center, Pax Axell Karamage, Rwanda Biomedical Center, Lawrence Ofori-Boadu, Ghana Health Service, Moses Mukiibi, Uganda Ministry of Health, Wesley Tomnos, Kenya Ministry of Health, Nyuma Mbewe, Zambia National Public Health Institute, Ijeoma Okoli, Federal Ministry of Health, Nigeria, Norman Lufesi, Malawi Ministry of Health.

## Notes

### Competing Interest Statement

Christian Ramers served on the Advisory Board for Pfizer and received consulting fees he has also received consulting fees or honoraria from Gilead Sciences, Viiv, and AbbVie. All other authors declare no competing interests

### Clinical Protocols

https://doi.org/10.1101/2024.10.25.24316111

### Funding Statement

This work was funded by grants from Pfizer, the Open Society Foundations, and the Conrad N. Hilton Foundation. The funders played no role in study design, data collection, analysis, or interpretation.

### Author Declarations

This study was approved by the Duke University Institutional Review Board (Pro00111388), The study was also approved by the local institutional review boards in each country participating in individual-level data collection: Ghana, Malawi, Rwanda, Zambia, and Nigeria.

